# Epigenetic dysregulation of *ACE2* and interferon-regulated genes might suggest increased COVID-19 susceptibility and severity in lupus patients

**DOI:** 10.1101/2020.03.30.20047852

**Authors:** Amr H. Sawalha, Ming Zhao, Patrick Coit, Qianjin Lu

## Abstract

Infection caused by SARS-CoV-2 can result in severe respiratory complications and death. Patients with a compromised immune system are expected to be more susceptible to a severe disease course. In this report we suggest that patients with systemic lupus erythematous might be especially prone to severe COVID-19 independent of their immunosuppressed state from lupus treatment. Specially, we provide evidence in lupus to suggest hypomethylation and overexpression of *ACE2*, which is located on the X chromosome and encodes a functional receptor for the SARS-CoV-2 spike glycoprotein. Oxidative stress induced by viral infections exacerbates the DNA methylation defect in lupus, possibly resulting in further *ACE2* hypomethylation and enhanced viremia. In addition, demethylation of interferon-regulated genes, NFκB, and key cytokine genes in lupus patients might exacerbate the immune response to SARS-CoV-2 and increase the likelihood of cytokine storm. These arguments suggest that inherent epigenetic dysregulation in lupus might facilitate viral entry, viremia, and an excessive immune response to SARS-CoV-2. Further, maintaining disease remission in lupus patients is critical to prevent a vicious cycle of demethylation and increased oxidative stress, which will exacerbate susceptibility to SARS-CoV-2 infection during the current pandemic. Epigenetic control of the *ACE2* gene might be a target for prevention and therapy in COVID-19.

---

Coronavirus Disease-2019 (COVID-19) is a respiratory disease caused by the severe acute respiratory syndrome coronavirus 2 (SARS-CoV-2) which is a novel single-stranded RNA virus of the *Coronaviridae* family. The disease can be associated with severe lower respiratory manifestations leading to acute respiratory distress syndrome, cytokine storm, and death ^1^. Older individuals and patients with pre-existing chronic medical illnesses appear to be at increased risk of severe COVID-19 ^2^.

Lupus is a chronic autoimmune disease of incompletely understood etiology. We propose that lupus patients might be at an increased risk for infection by SARS-CoV-2 and for developing a more severe form of COVID-19, independent of the possible effect of immunosuppressive medications.

Epigenetic dysregulation plays an important role in the pathogenesis of lupus ^3^. While most epigenetic studies in lupus have been performed in T cells, there is evidence to suggest reduced DNA methylation levels in multiple cell types in this disease. For example, a robust demethylation signature in interferon-regulated genes in lupus is wide-spread and characteristic of the disease ^4-6^.

DNA methylation plays an important role in the inactivation of the X chromosome. X chromosome inactivation, whereby gene expression from one gene copy of the X chromosome is silenced by DNA methylation, is essential to maintain normal levels of gene expression and comparable expression levels between male and female cells. In women with lupus, reduced DNA methylation has been shown to result in defective X chromosome inactivation ^7^. Hypomethylation of the normally inactivate X chromosome leads to an abnormal increase in the expression levels of affected genes. This abnormal X chromosome inactivation has been demonstrated to mediate increased expression of CD40LG in CD4+ T cells in women with lupus, contributing to T cell autoreactivity in this disease ^7^. CD40LG expression in CD4^+^ T cells isolated from healthy women was similarly increased after treatment with the DNA methylation inhibitor 5-azacytidine. This gene-dose effect of the X chromosome has been proposed to explain, at least in part, sex bias in lupus. Of interest, men with two X chromosomes, Klinefelter syndrome (47, XXY), have a significantly higher risk to develop lupus compared to normal men (46, XY) and a similar risk to women ^8^.

SARS-CoV-2 binds to target host cells through angiotensin-converting enzyme 2 (ACE2). ACE2 is a functional receptor for the viral spike glycoprotein that allows the entry of SARS-CoV-2 into cells ^9^. *ACE2* gene is located on the X chromosome, similar to *CD40LG*. Recent findings suggest the expansion of a demethylated T cells subset in lupus patients characterized by the expression of several genes known to be demethylated in lupus, including *CD40LG, CD70, ITGAL* (*CD11A*), and the KIR gene cluster (defined using flow cytometry as KIR^+^CD11a^hi^ CD4^+^CD28^+^ T cell subset) ^10,11^. Indeed, we can induce this T cell subset in normal healthy individuals using the DNA methylation inhibitor 5-azacytidine ^11^. We generated whole-genome DNA methylation data using an array-based approach and observe significant hypomethylation in the *ACE2* gene in this T cell subset compared to KIR^-^CD11a^low^ T cells isolated from the same lupus patients (**Figure 1A**). This hypomethylation involves several CpG sites located in the *ACE2* promoter region proximal to the transcription start site, the 5’-untranslated region, and the 3’-untranslated region, suggesting functional regulatory effect of the methylation changes. Furthermore, we can induce hypomethylation in some of these same CpG sites in CD4^+^ T cells isolated from normal healthy women treated with the DNA methylation inhibitor 5-azacytidine ^11^. Importantly, in a subset of female lupus patients characterized by skin, joint, and kidney involvement *ACE2* promoter region was significantly hypomethylated in CD4^+^ T cells compared to age and sex matched normal healthy controls^12^. Gene expression profiles extracted from a publicly available dataset (GEO accession: GSE4588) showed a significant overexpression of ACE2 mRNA in lupus CD4^+^ T cells compared to normal healthy controls (**Figure 1B**). We further confirm hypomethylation in *ACE2* in an independent set of female lupus patients characterized by kidney involvement (**Figure 1C**). Gene expression profiles in the same set of patients showed ∼1.5-2-fold ACE2 overexpression compared to healthy controls, although his did not reach statistical significance (**Figure 1D**).

**Figure 1:**
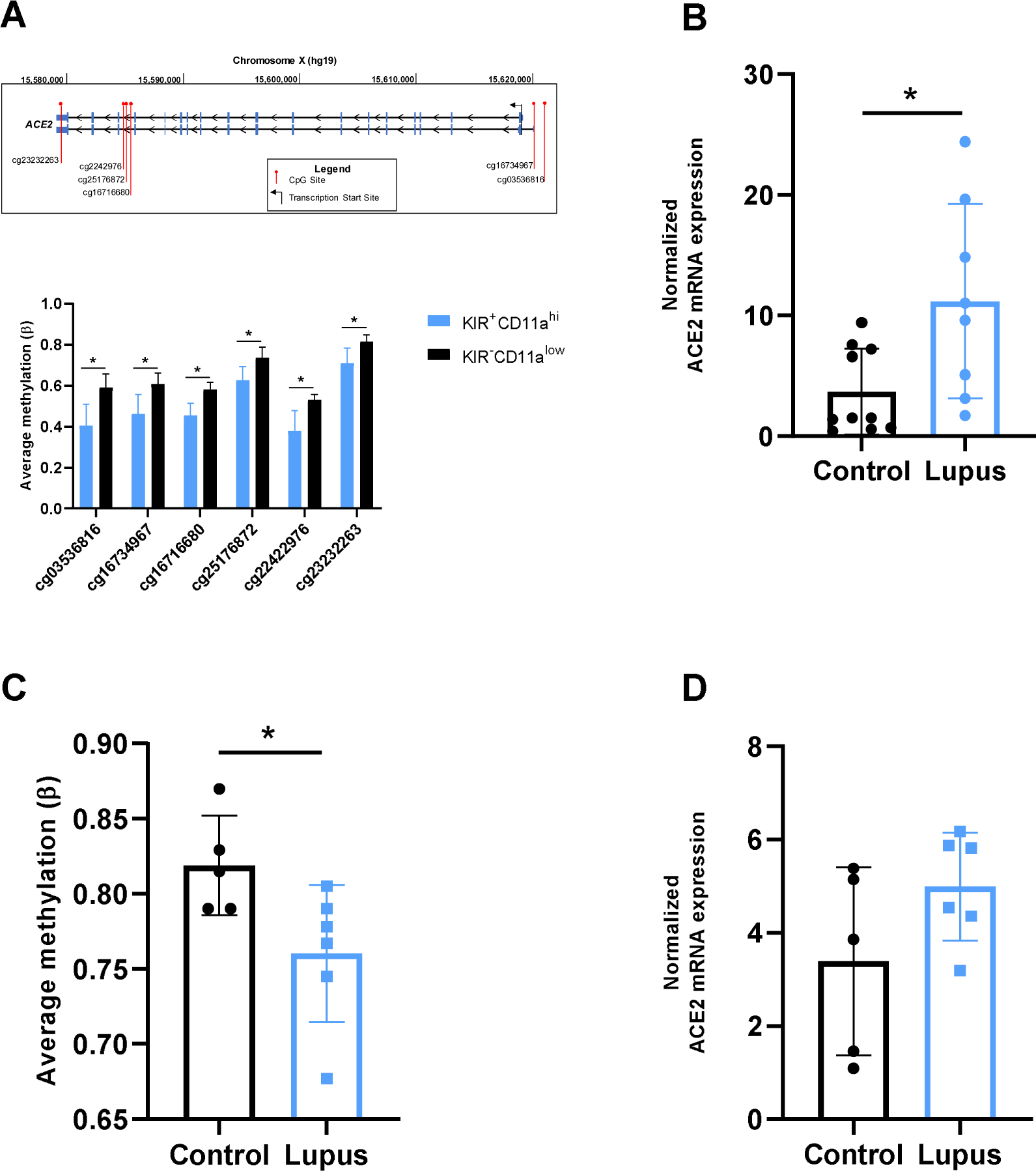
**A)** DNA hypomethylation of *ACE2* gene in a subset of CD4^+^ T cells characterized by expression of KIR genes and high levels of CD11a (KIR^+^CD11a^hi^) compared to autologous KIR^-^CD11a^low^ CD4^+^ T cells in lupus patients. The KIR^+^CD11a^hi^ T cell subset has been previously shown to overexpress other methylation sensitive genes in lupus patients including *CD40LG*, also located on the X chromosome. (*, P< 0.01, corrected for multiple testing and derived using GenomeStudio (Illumina)). **B)** Normalized mRNA expression values of ACE2 in CD4^+^ T cells isolated from lupus patients compared to healthy controls. These data were extracted from Gene Expression Omnibus (GEO accession: GSE4588) (P= 0.01; Mann-Whitney test). **C)** Demethylation of a CpG site in the 3’-untranslated region of *ACE2* (cg23232263) in CD4+ T cells from lupus patients with a history of kidney involvement compared to age, sex, and ethnicity matched healthy controls. DNA methylation beta values were extracted from genome-wide methylation data generated using the HumanMethylation450 array (P= 0.03; Mann-Whitney test) **D)** Normalized mRNA expression values of ACE2 in the same CD4+ T cell samples from lupus patients and controls shown in panel C. Values were extracted from Affymetrix U133 plus 2.0 array data (P= 0.18; Mann-Whitney test).

Lupus patients are also characterized by expansion of a pro-inflammatory double negative T cell subset (CD4^-^CD8^-^) ^13^. We have previously demonstrated that double negative T cells heavily demethylate cytokine genes including IL-18 and IFNγ, among others ^14^. Interestingly, we also observe significant hypomethylation in the *ACE2* gene in double negative T cells compared to autologous CD4+ and CD8+ T cells isolated from normal healthy women ^14^.

Together, these data support that ACE2 expression is regulated by DNA methylation and that a methylation defect in lupus extends to regulatory sequences in the *ACE2* gene, which could result in ACE overexpression in lupus patients.

It is important to note that a major limitation to these arguments is the absence of data from alveolar epithelial cells in lupus patients. However, it is worth highlighting that overexpression of ACE2 in other cell types might facilitate viremia and organ damage in COVID-19. Indeed, in SARS, SARS-CoV viral particles (which also utilize ACE2 for cell entry) have been shown to infect immune cells in large numbers, including peripheral blood T cells, which can result in viral dissemination throughout the body ^15^. Direct tissue damage from viral infection of target tissues, such as the kidneys, intestine, and brain has been reported ^16^.

Overall, the DNA methylation defect in lupus patients is more evident in patients with increased disease activity, and is exacerbated by increased oxidative stress, such as in the case of viral infections ^17^. Oxidative stress has been shown to inhibit DNA methylation in lupus T cells by blocking PKC-δ activation, resulting in attenuated MEK/ERK signaling and reduced expression of the main DNA methyltransferase DNMT1 ^17,18^. In addition, oxidative stress depletes NADPH and glutathione levels leading to mTOR activation which also exacerbates the DNA methylation defect by inhibiting DNMT1 ^19^. Therefore, it is reasonable to suggest the possibility that the DNA methylation defect in lupus patients, exacerbated by the oxidative stress from SARS-CoV-2 infection, will further enhance viral entry in lupus patients through epigenetic de-repression of *ACE2* and increased ACE2 expression.

Lupus patients are characterized by robust demethylation of interferon-regulated genes, which is persistent and present across multiple cells types and disease subsets ^20^. We have previously suggested that this epigenetic activation poises interferon-regulated genes for transcription in lupus patients, and likely explains the exaggerated expression response of interferon-regulated genes in the presence of response to type-I interferons ^4^.Therefore, it is possible that an interferon response induced by viral infections, including SARS-CoV-2, is more likely to be complicated by an exaggerated immune-response in lupus patients. This might contribute to a cytokine storm and tissue damage during COVID-19 infections in lupus patients. Indeed, we have also demonstrated progressive demethylation in CD4^+^ T cells of the gene that encodes the master cytokine regulator NFκB and a number of pro-inflammatory cytokine genes and their key transcription factors with increasing disease activity in lupus ^21^. Response to SARS-CoV-2 resulting in tissue damage and oxidative stress might further exacerbate the pro-inflammatory epigenetic changes observed in lupus patients resulting in a vicious circle of cytokine response (**Figure 2**).

**Figure 2:**
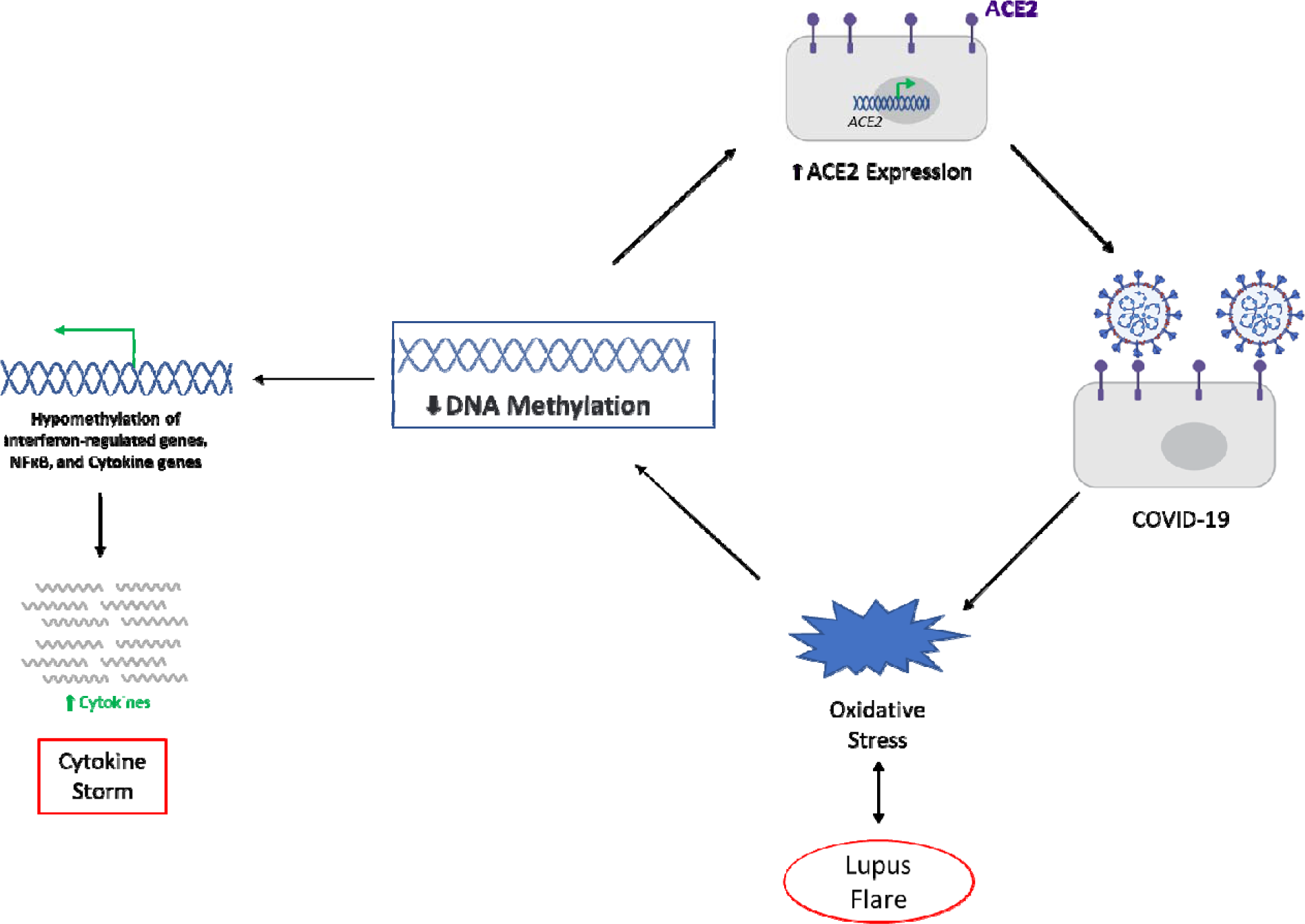
Schematic presentation suggesting mechanisms that might increase susceptibility to and severity of COVID-19 in lupus patients. Lupus is characterized by a DNA methylation defect which has been demonstrated in multiple cell types. SARS-CoV-2 utilizes ACE2 as a receptor for entry into cells. *ACE2* is a methylation-sensitive gene located on the X chromosome. Lupus patients show evidence of *ACE2* hypomethylation possibly mediating ACE2 overexpression which will increase the likelihood of SARS-CoV-2 infection. Viral infections are associated with increased oxidative stress, which is known to induce lupus flares and induce further DNA demethylation. Hypomethylation and overexpression of *ACE2* in T cells (and possibly other cell types) will facilitate T cell viral infections and viral dissemination resulting in a more severe COVID-19. Interferon-regulated genes and other inflammatory cytokine genes are hypomethylated in lupus patients and thereby epigenetically poised or primed for transcription upon interferon exposure resulting from viral immune response. This epigenetic priming might increase the possibility of cytokine storm in lupus patients.

Taken together, these observations suggest that epigenetic dysregulation might increase the risk and severity of SARS-CoV-2 infections in lupus patients. The likelihood of disseminated SARS-CoV-2 infection in lupus patients might be increased due to ACE2 overexpression in peripheral blood mononuclear cells. This is independent of any potential effects of immunosuppressive medications. Based on this model, we also expect that lupus flares will increase the likelihood of infection with SARS-CoV-2 due to increased oxidative stress and DNA demethylation of *ACE2*. Therefore, maintaining remission in lupus patients is of critical importance during this pandemic. Drugs that target epigenetic mechanisms should be considered for further investigation in COVID-19.

## Data Availability

All data are available within the manuscript

## Ethical approval information

All study participants signed informed consents approved by the institutional review boards/ethics committees of our institutions.

